# Gut microbiota alterations in patients with persistent respiratory dysfunction three months after severe COVID-19

**DOI:** 10.1101/2021.07.13.21260412

**Authors:** Beate Vestad, Thor Ueland, Tøri Vigeland Lerum, Tuva Børresdatter Dahl, Kristian Holm, Andreas Barratt-Due, Trine Kåsine, Anne Ma Dyrhol-Riise, Birgitte Stiksrud, Kristian Tonby, Hedda Hoel, Inge Christoffer Olsen, Katerina Nezvalova Henriksen, Anders Tveita, Ravinea Manotheepan, Mette Haugli, Ragnhild Eiken, Åse Berg, Bente Halvorsen, Tove Lekva, Trine Ranheim, Annika Elisabeth Michelsen, Anders Benjamin Kildal, Asgeir Johannessen, Lars Thoresen, Hilde Skudal, Bård Reiakvam Kittang, Roy Bjørkholt Olsen, Carl Magnus Ystrøm, Nina Vibeche Skei, Raisa Hannula, Saad Aballi, Reidar Kvåle, Ole Henning Skjønsberg, Pål Aukrust, Johannes Roksund Hov, Marius Trøseid, on behalf of the NOR-Solidarity study group

## Abstract

**Objective:** Although COVID-19 is primarily a respiratory infection, mounting evidence suggests that the GI tract is involved in the disease, with gut barrier dysfunction and gut microbiota alterations being related to disease severity. Whether these alterations persist and could be related to long-term respiratory dysfunction is unknown.

**Design:** From the NOR-Solidarity trial (n=181), plasma was collected during hospital admission and after three months, and analyzed for markers of gut barrier dysfunction and inflammation. At the three-month follow-up, pulmonary function was assessed by measuring diffusing capacity of the lungs for carbon monoxide (DL_CO_), and rectal swabs for gut microbiota analyses were collected (n= 97) and analysed by sequencing of the 16S rRNA gene.

**Results:** Gut microbiota diversity was reduced in COVID-19 patients with persistent respiratory dysfunction, defined as DL_CO_ below lower limit of normal three months after hospitalization. These patients also had an altered global gut microbiota composition, with reduced abundance of *Erysipelotrichaceae UCG-003* and increased abundance of *Flavonifractor* and *Veillonella*, the latter potentially being linked to fibrosis. During hospitalization, increased plasma levels of lipopolysaccharide-binding protein (LBP) were strongly associated with respiratory failure, defined as pO_2_/fiO_2_-(P/F-ratio)<26.6 kPa. LBP levels remained elevated during and after hospitalization, and was associated with low-grade inflammation and persistent respiratory dysfunction after three months.

**Conclusion:** Persistent respiratory dysfunction after COVID-19 is associated with reduced biodiversity and gut microbiota alterations, along with persistently elevated LBP levels. Our results point to a potential gut-lung axis that should be further investigated in relation to long-term pulmonary dysfunction and long COVID.

**Summary box:** *What is already known about this subject?:*

- Mounting evidence suggests that the gastrointestinal tract is involved in the pathogenesis of COVID-19, with the putative SARS-CoV-2 receptor ACE 2 ubiquitously expressed in the gut.
- In severe COVID-19, the gut-blood barrier is compromised, and leakage of microbial products, such as lipopolysaccharides (LPS), could affect the host’s response to COVID-19 infection.
- COVID-19 patients exhibit an altered gut microbiota composition, which has been related to disease severity. However, it is currently not known whether dysbiosis or gut barrier dysfunction persist long-term after hospitalization, or whether microbiota-related mechanisms could be related to persistent pulmonary dysfunction.

*What are the new findings?:*

- COVID-19 patients with persistent respiratory dysfunction after three months had a lower microbial diversity and an altered gut microbiota composition at the same time point.
- The microbiota alterations included reduced abundance of *Erysipelotrichaceae UCG-003* and increased abundance of *Veillonella* and *Flavonifractor*.
- During hospitalization, increased plasma levels of LBP were strongly associated with respiratory failure.
- LBP levels remained elevated during and after hospitalization, and associated significantly with persistent respiratory dysfunction at three-month follow-up.

*How might it impact on clinical practice in the foreseeable future?:* Our findings point to a potential gut-lung axis in relation not only to respiratory failure during hospitalization, but also to long-term COVID-19 morbidity. Further studies on gut microbiota composition and gut barrier dysfunction as potential treatment targets and/or disease severity biomarkers in relation to long-term pulmonary dysfunction and long COVID are warranted.

## Introduction

Although COVID-19 is primarily a viral respiratory disease, aberrant immune responses to the causative virus, SARS-CoV-2, resulting in systemic inflammation and multi-organ involvement are central features in severe and critical disease. In addition, mounting evidence suggests that the gastrointestinal (GI) tract is involved in the pathogenesis of COVID-19(1, 2). This includes the ability of SARS-CoV-2 to infect and replicate within human small intestine enterocytes, with viral RNA being detectable in fecal samples long time after symptom relief(3, 4).

The GI tract is the largest immunological organ in the body, and its resident microbiota is known to modulate regional as well as systemic host immune responses. It has been hypothesized that the gut microbiota could be a mediator of host inflammatory immune responses during COVID-19 and thereby contribute to the pronounced systemic inflammation seen in patients requiring hospitalization(5, 6). In fact, angiotensin-converting enzyme (ACE) 2, the putative SARS-CoV-2 receptor, is ubiquitously expressed in several organs, including the gut, where ACE2 expression in enterocytes may serve as a site for enteric SARS-CoV-2 infection(7).

Recent studies suggest that severe SARS-CoV-2 infection could compromise the integrity of the gut-blood barrier, leading to enhanced leakage of microbial products such as lipopolysaccharides (LPS), possibly affecting the host’s response to COVID-19 through activation of the innate immune system(3, 8). In a small observational study of hospitalized COVID-19 patients, we recently reported that elevated plasma levels of LPS-binding protein (LBP) were associated with elevated cardiac markers(7). We also found elevated levels of the CCR9-ligand CCL25 in these patients, which is involved in gut homing of T cells(9). However, knowledge on the relationship between markers of gut barrier dysfunction and the degree of respiratory failure, and in particular persistently impaired pulmonary dysfunction following COVID-19 hospitalization, is scarce.

COVID-19 patients exhibit an altered gut microbiota composition(3, 4), and gut microbiome alterations were recently found to be related to disease severity(10). Although these alterations persisted when investigated at median six days after a negative SARS-CoV-2 PCR test, it is currently not known whether gut microbiota alterations or gut barrier dysfunction persist long-term after hospitalization. Moreover, whether microbiota-related mechanisms could be involved in the persistent inflammation and pulmonary dysfunction observed in some patients at follow-up after hospitalization for COVID-19 is unknown.

The NOR-Solidarity trial is an independent add-on study to the WHO Solidarity trial(11), recently reporting no effects of hydroxychloroquine (HCQ) or remdesivir compared to standard of care (SoC) on clinical outcome, viral clearance or systemic inflammation in hospitalized COVID-19 patients(12). In the present sub-study, we investigated whether gut microbiota collected three months after hospital admission, as well markers of gut barrier dysfunction measured during and after hospitalization, were related to persistent pulmonary dysfunction after COVID-19.

## Methods

### Study design and participants

NOR-Solidarity is a multicentre, open label, adaptive randomised clinical trial evaluating the effect of antiviral drugs on hospitalized COVID-19 patients admitted to 23 Norwegian hospitals(12). In addition, NOR-Solidarity included collections of blood biobank and outpatient visits at a three-month follow-up after hospital admission. The study was approved by the Committee for Medical Research Ethics Region South East Norway (118684) and by the Norwegian Medicines Agency (20/04950-23) and registered in ClinicalTrials.gov (NCT04321616). Participants were included from March 28^th^ until October 5^th^ 2020, and all participants >18 years admitted to the hospital with PCR-confirmed SARS-2-CoV-2 infection were eligible for inclusion. Exclusion criteria are described in the original study protocol and included severe comorbidity (life expectancy < 3 months), high levels of liver transaminases (AST/ALT > 5 times upper limit of normal), corrected QT interval time as assessed by ECG more than 470 ms, pregnancy, breast feeding, acute comorbidity occurrence in a 7-day period before inclusion, known intolerance to study drugs, concomitant medications interfering with the study drugs or participation in a confounding trial(12). All participants gave informed consent prior to inclusion, either by themselves or a legally authorised representative.

### Intervention and outcomes

In the NOR-Solidarity study (n=181), participants were randomized and allocated to one of three treatment arms, 1) local SoC; 2) SoC plus 800 mg of oral HCQ twice daily on day 1, then 400 mg twice daily up to 9 days; or 3) SoC plus 200 mg of intravenous remdesivir on day 1, then 100 mg daily up to 9 days. All study treatments were stopped at hospital discharge. Since the interventions had no effect on clinical outcome, viral clearance or systemic inflammation(12), data from the different intervention arms were in this sub-study pooled for analyses to examine whether gut microbiota composition after three months and markers of gut barrier dysfunction during hospitalization and after three months had any effect on:(i) acute respiratory failure defined as pO_2_/fiO_2_-(P/F-ratio) <26.6 kPa (<200 mmHg) during hospitalization, (ii) persistent respiratory dysfunction defined as diffusing capacity of the lungs for carbon monoxide (DL_CO_) below the lower limit of normal (LLN) at the three-month follow-up. To ensure lack of influence of different treatment, data were also adjusted for treatment group.

### Circulating markers of gut barrier dysfunction

Peripheral blood samples were drawn at admission, day three and day eight and then weekly until hospital discharge, as well as at outpatient follow-up at three months. Age- and sex-matched controls (Regional Committees for Medical and Health Research 2014/2078) were used as reference material. Plasma levels of LBP, intestinal fatty acid binding protein (IFABP), CCL25 and regenerating islet-derived protein 3α (REG-3α) were measured in duplicate by enzyme immunoassays (EIA) using commercially available antibodies (R&D Systems, Minneapolis, MN) in a 384 format using a combination of a SELMA (Jena, Germany) pipetting robot and a BioTek (Winooski, VT) dispenser/washer. Absorption was read at 450 nm with wavelength correction set to 540 nm using an EIA plate reader (Bio-Rad, Hercules, CA). Intra- and inter-assay coefficients of variation were <10%.

### Three-month follow-up

Three months after hospital admission, 149 participants attended a follow-up visit that included blood sampling for routine clinical biochemistry and biobanking(12). Rectal swabs were also collected from a sub-group of participants (n=97). Lung function tests (n=108), consisting of spirometry and diffusion capacity of the lungs for carbon monoxide (DL_CO_), were performed at each participating center, and performed according to the European Respiratory Society’s and the American Thoracic Society’a guidelines(13, 14), thoroughly described previously(15). Studies have shown that DL_CO_ is the pulmonary measure most frequently affected after hospitalization for COVID-19(15, 16), and for the purpose of this report, we chose DL_CO_ as a measure of pulmonary function. DL_CO_ in per cent of predicted and the lower limit of normal (LLN) were calculated according to the Global Lung Function Initiative Network (GLI)(17), as previously described(15). Of the 108 participants that underwent lung function tests, 83 participants also had rectal swabs available.

### Gut microbiota analyses

Rectal swabs were stored on a stabilizing transportation medium (soluble Amies, Thermo Scientific(tm)) and frozen at -80C until analysed. Fecal DNA was extracted using the QIAamp® PowerFecal®Pro DNA Kit (Qiagen, Germany), with slight modifications. Briefly, 700 µL of fecal material was pelleted and homogenized in 800 µL of kit solution CD1 using a bead-beater (2×60s at 5.5 ms, room temperature), and further processed according to the manufacturer’s protocol. Libraries for 16S rRNA amplicon sequencing were generated as previously described(18). Briefly, the hypervariable regions V3 and V4 of the 16S rRNA gene were amplified using dual-indexed universal primers 319F (forward) and 806R (reverse) and Phusion High-Fidelity PCR Master mix with HF buffer (Thermo Fisher Scientific, USA). Cleaning and normalization of PCR products were performed using the SequalPrep Normalization Plate Kit (Thermo Fisher Scientific, USA). Quality control and quantification of pooled libraries were performed using Agilent Bioanalyzer (Agilent Technologies, USA) and Kapa Library Quantification Kit (Kapa Biosystems, London, UK). Sequencing was performed at the Norwegian Sequencing Centre (Oslo, Norway), applying the Illumina MiSeq platform and v3 kit (Illumina, San Diego, CA, USA), allowing for 300bp paired-end reads.

### Sequence processing and bioinformatics

Paired-end reads containing Illumina Universal Adapters or PhiX were discarded using bbduk version 38.90 (BBTools, https://jgi.doe.gov/data-and-tools/bbtools/) (parameters adaptor filter: k=23 hdist=1 tbo cf=TRUE ftm=5. parameters phix filter: k=31 hdist=1) and the remaining reads were demultiplexed using cutadapt version 3.3(19) (parameters:-e 0.1 --no-indels --overlap 12 --discard-untrimmed --action none). Trimming of indexes, heterogeneity spacers and primers was also done with cutadapt (parameters: -e 0.1 --overlap 20 --discard-untrimmed -m 250) and the paired-end reads were subsequently quality trimmed and merged using bbmerge version 38.90(20) (parameters: qtrim=r trimq=15 maxlength=440 mininsert=390). The merged contigs were trimmed to 400bp and denoised to ASVs (Amplicon Sequence Variants) with deblur(21) in QIIME2 version 2021.2(22). Taxonomic classification of ASVs was done based on RESCRIPt(23) in QIIME2 using a naïve Bayes classifier trained on the V3-V4 region of a preclustered version (99% sequence similarity) of the Silva database version 138(24). Filtering of contaminants was done with the R package microDecon(25) based on a negative extraction control sample, and ASVs from mitochondria, chloroplast or lacking taxonomic annotation on order level were also removed. A de-novo phylogenetic tree was built in QIIME2 based on the remaining ASVs. To reduce the effect of uneven sequencing depths, samples were rarefied (subsampled without replacement) to an even level of 7123 counts per sample and all diversity analyses were performed on this rarefied dataset. Alpha diversity metrics (Observed ASVs and Faith’s phylogenetic diversity, PD) and beta diversity metrics (Bray-Curtis dissimilarity) were calculated in QIIME2. Beta diversity comparisons were done with a permutational multivariate analysis of variance test (Permanova). Differential abundance testing with LEfSe(26) was done on the rarefied dataset, while for differential abundance testing with ALDEx2(27), a prevalence-filtered (25% most prevalent) version of the non-rarefied dataset was used.

### Statistical analyses

Baseline characteristics were described by median (25th and 75th percentile (IQR)) for continuous variables and percentages for categorical variables. Predefined outcomes (acute respiratory failure defined as P/F-ratio <26.6 kPa during hospitalization and persistent respiratory dysfunction defined as DL_CO_<LLN after 3 months follow-up) were used as dichotomous variables. Analysis of clinical characteristics at hospital admission (Fig. 4) according to acute respiratory failure was analyzed by receiver operating characteristic (ROC) analysis and precision-recall curves were assessed, as the outcome groups were unbalanced. IFABP, CCL25 and LBP were non-normally distributed and transformed using log10 for comparisons between groups with the linear mixed model analysis, with subject as random effect and time and respiratory failure as fixed effects (also as interaction). Treatment was included as a covariate. Multivariate analysis of LBP in relation to respiratory failure was performed by binary logistic regression, adjusting for the covariates age, gender, treatment and known comorbidities in the base model. Correlation analyses were performed with Spearman’s rho (ρ) due to skewed distribution of data. P-values are two-sided and considered significant when <0.05. Group comparison of LBP levels in relation to persistent respiratory dysfunction was performed using parametric t-test of log10-transformed data. SPSS release 26.0.0.1 and 27.0.0.0 were used for statistical analysis.

## Results

### Baseline characteristics

NOR-Solidarity trial design and main results have recently been published(12). A total of 181 randomized patients were included in the present study and 149 completed the three months follow-up, of which 108 performed pulmonary function testing and 97 had collected rectal swab for gut microbiota analyses (Fig. 1). Baseline characteristics for the main study group and for the microbiota sub-group are given in Table 1. In the main study group, participants were on average 59 years, mostly male (66%), and the median body mass index (BMI) was 27.4 kg/m^2^ (27% obese, BMI > 30 kg/m^2^). Use of antibiotics during hospitalization was reported in 48% of the participants, and 68% reported any known comorbidities, with hypertension (31%), obesity (27%) and diabetes mellitus (17%) as the most common. Only six per cent reported presence of chronic pulmonary disease.

**Table 1.** Baseline characteristics.

|  | Main study group<br>(n=181) | Microbiota sub-group<br>(n=97) |
| --- | --- | --- |
| Age, years | 59 (50.0-71.0) | 57 (48.0-65.0) |
| Male gender (%) | 119 (66) | 60 (62) |
| Body Mass Index, kg/m<sup>2</sup> | 27.4 (24.7-30.9) | 27.7 (25.0-31.6) |
| Antibiotics use (%) | 87 (48) | 41 (42) |
| Treatment group |  |  |
| Standard of Care (SoC) (%) | 87 (48) | 47 (49) |
| SoC + Hydroxychloroquine (%) | 52 (29) | 33 (34) |
| SoC + Remdesivir (%) | 42 (23) | 17 (18) |
| Comorbidities |  |  |
| Any known comorbidities (%) | 122 (68) | 66 (68) |
| Chronic pulmonary disease (%) | 10 (6) | 5 (5) |
| Hypertension (%) | 55 (31) | 28 (29) |
| Chronic cardiac disease (%) | 28 (16) | 12 (12) |
| Diabetes mellitus (%) | 31 (17) | 19 (20) |
| Obesity, BMI > 30 kg/m<sup>2</sup> (%) | 44 (27) | 28 (30) |
| P/F-ratio at admission, kPa | 42.1 (32.0-48.1) | 42.4 (33.4-49.6) |
| C-reactive protein, mg/L | 70.0 (36.3-70.0) | 66.0 (33.5-122.5) |
| White blood cell count, x 10<sup>9</sup>/L | 6.2 (4.7-8.6) | 5.8 (4.3-7.6) |
| Neutrophil count, x10<sup>9</sup>/L | 4.3 (3.0-6.6) | 3.7 (2.4-6.2) |
| Ferritin, µg/L | 626 (325-1209) | 608 (298-1017) |
| D-dimer, mg/L | 0.7 (0.5-1.2) | 0.7 (0.4-1.1) |
| Virus load (log<sub>10</sub>/1000) | 1.9 (0.7-3.1) | 1.7 (0.0-2.9) |
| Lipopolysaccharide-binding protein (µg/mL) | 16.3 (8.5-26.1) | 17.1 (7.3-26.3) |

Results are shown as median values (25th-75th percentile) unless otherwise specified.

**Figure 1.**
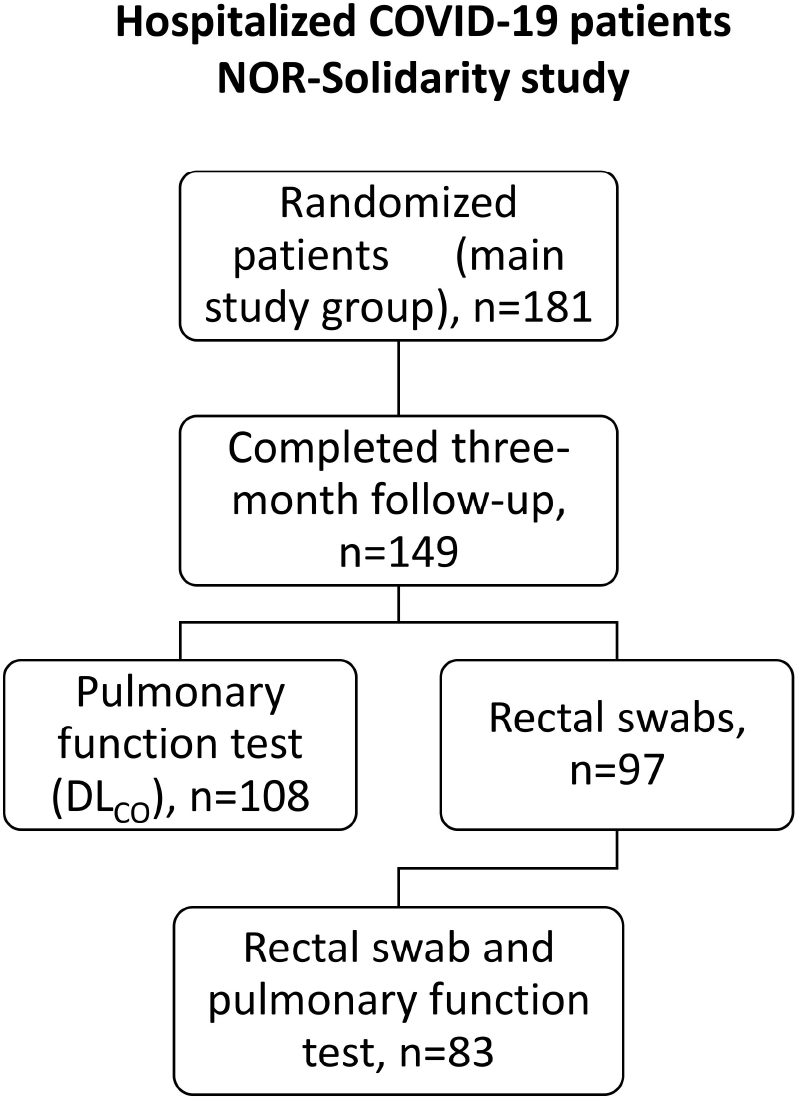
Flow-chart of patients included in the present add-on study based on the original NOR-Solidarity study protocol.

### Respiratory dysfunction after three months is associated with altered gut microbiota composition and reduced bacterial diversity in previously hospitalized COVID-19 patients

At the three-month follow-up, 25 out of the 83 (30%) previously hospitalized COVID-19 patients with rectal swabs and DL_CO_ measurements available, had respiratory dysfunction defined as DL_CO_<LLN. Notably, these patients had an altered global gut microbiota composition assessed by beta diversity (Bray Curtis (Fig. 2A)). Importantly, patients who received antibiotics during hospitalization had overlapping global microbiota composition with those not receiving antibiotics (Fig. 2B). Moreover, there was no statistical interaction between antibiotics use during hospitalization and the association between global microbiota composition and respiratory dysfunction at the three-month follow up.

**Figure 2.**
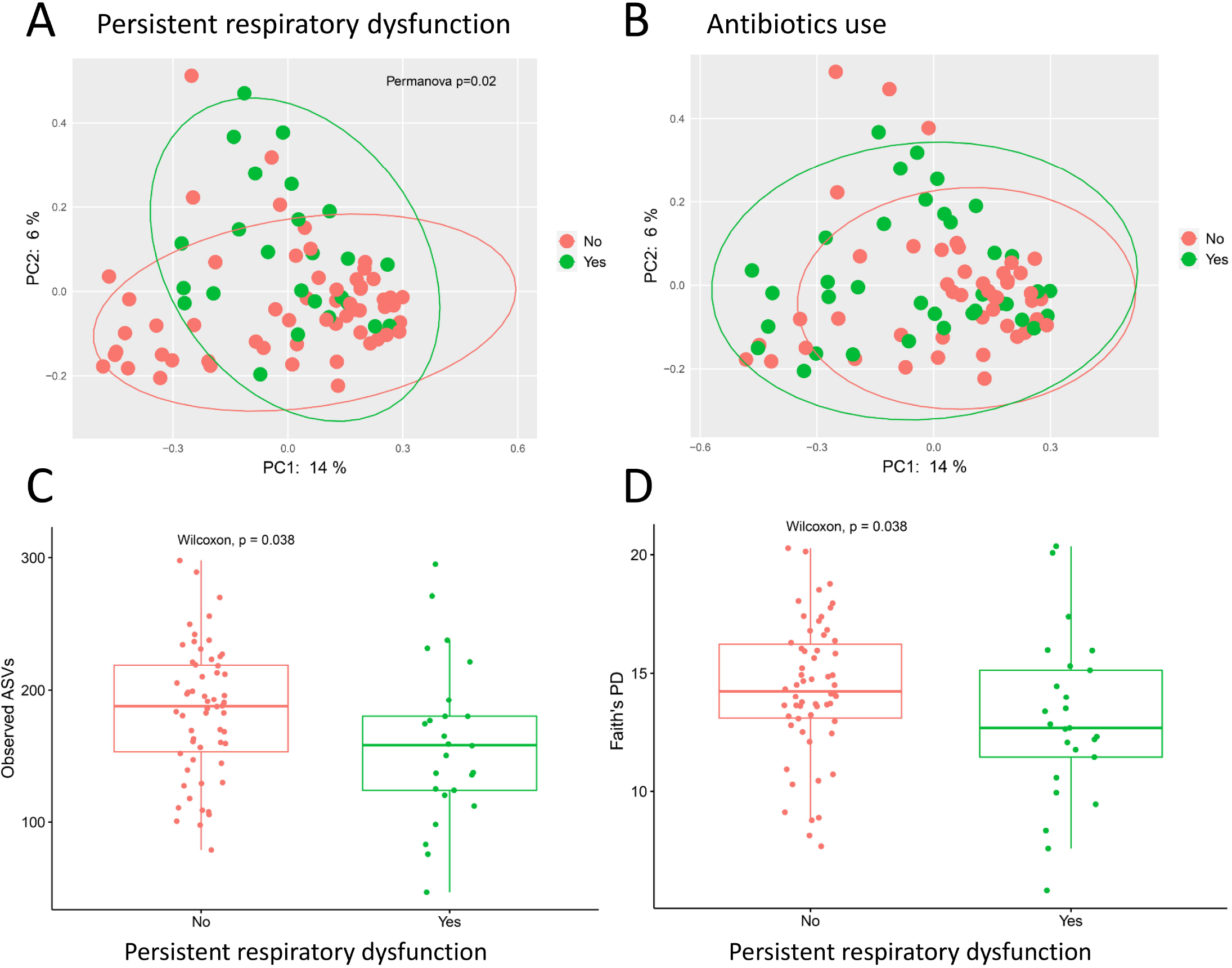
Gut microbiota diversity in patients with or without persistent respiratory dysfunction at the three-month follow-up. Beta diversity by principal coordinate analysis showing Bray-Curtis distances separating patients with or without respiratory dysfunction (DLCO<LLN, n=83) (A), with overlapping microbiota composition in relation to antibiotics use (B). Alpha diversity measured by observed ASVs (C) and Faith’s PD (D) in patients with or without respiratory dysfunction. Abbreviations: ASVs, amplicon sequence variants; PD, phylogenetic diversity.

Patients with respiratory dysfunction three months after hospital admission also had reduced bacterial alpha diversity (Observed ASVs and Faith’s PD) compared to patients with respiratory function in the normal range (Fig. 2C-D). However, alpha diversity measures did not associate with age, gender, comorbidities, BMI, treatment groups, viral load or antibiotics use.

On taxonomic level, screening analysis using LEfSe suggested that respiratory dysfunction after three months was associated with increased relative abundances of five taxa and reduced relative abundances of 20 taxa (Fig. 3A-B), including *Erysipelotrichaceae UCG-003*, and also several members of the Lachnospiraceae and Ruminococcaceae families, which are known producers of butyrate, the main energy source for enterocytes. We subsequently performed the same analysis using the ALDEx2 algorithm (filtered to p<0.05), reducing the number of genera identified by both LEfSe and ALDEx2 as associated with respiratory dysfunction to three; reduced abundance of *Erysipelotrichaceae UCG-003*, and increased abundance of *Veillonella* and *Flavonifractor* (Fig. 3C). The largest effect size and lowest p-value from ALDEx2 was found for *Veillonella*, an anaerobic opportunistic pathogen, previously reported increased in COVID-19 patients (4) and associated with tissue fibrosis(28).

**Figure 3.**
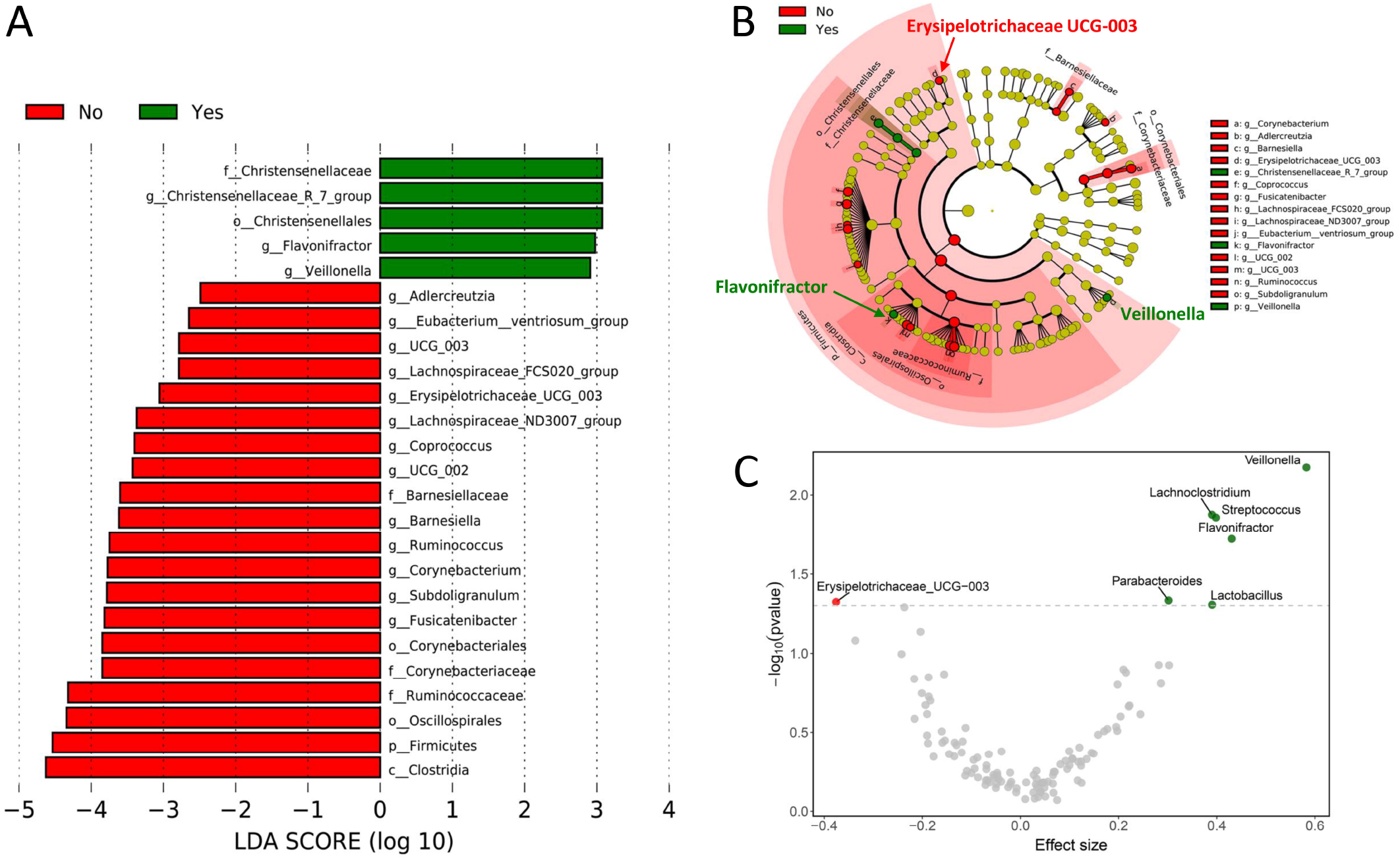
Gut microbial composition in patients with persistent respiratory dysfunction at the three-month follow-up (DLCO<LLN, n=83). A) LDA score of taxa abundance differences using LEfSe analysis. B) Taxonomic cladogram highlighting differentially abundant taxa (p<0.05) by LEfSe. C) Volcano plot from ALDEx2 analysis showing effect size (difference between groups divided by dispersion within groups, on log2-scale) by p-value of differentially abundant genera. Abbreviations: LDA, linear discriminant analysis; LEfSe, linear discriminant analysis effect size.

**Figure 4.**
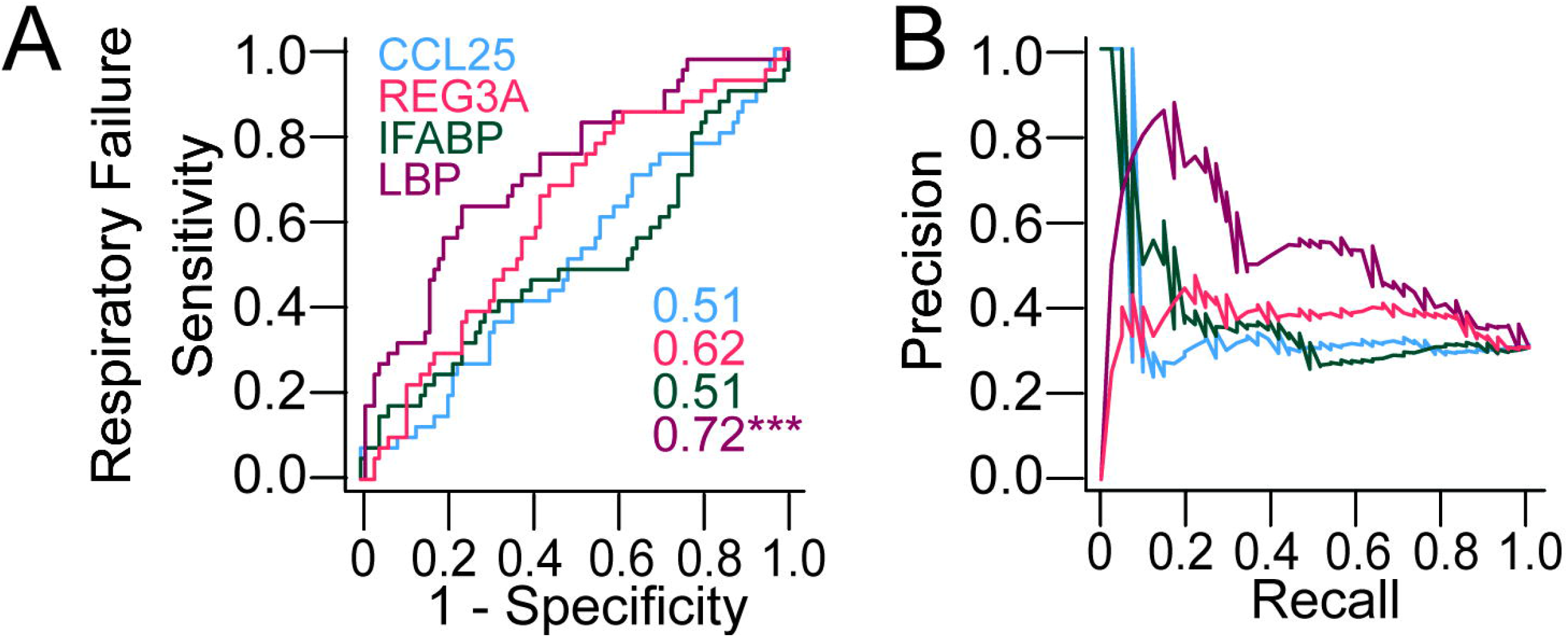
Admission levels (n=144) of markers of gut barrier dysfunction in relation to acute respiratory failure (n=44) during hospitalization (P/F-ratio <26.6 kPa). A) ROC analysis and B) precision-recall curves for LBP (brown), IFABP (green), CCL25 (blue) and REG-3α (orange). The numbers shown in A) are the AUC from the ROC-analysis ***p<0.001. Abbreviations: LBP, lipopolysaccharide-binding protein; IFABP, intestinal-type fatty acid-binding protein; CCL25, chemokine (C-C motif) ligand 25; REG-3α, regenerating islet-derived protein 3 alpha; ROC, receiver operating characteristic; AUC, area under the curve.

### Circulating markers of gut barrier dysfunction in relation to acute respiratory failure during hospitalization

As microbiota traits associated with persistent respiratory dysfunction were potentially related to reduced capacity for butyrate production, which is vital for a functional gut barrier, we hypothesized that gut barrier dysfunction could be related to respiratory failure also in acute COVID-19. As microbiota samples from the acute phase of disease were not available, we explored whether circulating markers of gut barrier dysfunction were associated with acute respiratory failure defined as P/F ratio <26.6 kPa, which occurred in 60 patients (33%) during hospitalization. As depicted in Fig. 4, levels of LBP discriminated clearly in ROC analyses (AUC 0.72), whereas CCL25, REG-3α and IFABP did not provide prognostic information (AUC between 0.62 and 0.51). However, as shown in the in the precison-recall curve (Fig. 4B), only around 60% (recall) of the patients with acute respiratory failure would be identified with only around 55% (precision) being true positives. In multivariate logistic regression adjusting for age, gender, comorbidity and treatment, LBP was associated with acute respiratory failure during hospitalization (adjusted OR (aOR) 6.3 [1.5-27]). Further adjusting for antibiotics use had little impact on the association (aOR 4.6 [1.0-21]).

### Temporal changes in markers of gut barrier dysfunction during hospitalization

LBP levels remained consistently elevated during hospitalization in patients with acute respiratory failure compared to patients without respiratory failure (Fig. 5A). However, whereas LBP levels showed a declining trend over time, the opposite was observed for REG-3α (Fig. 5D), a bactericidal C-type lectin with protective antibacterial properties, produced in the intestine. REG-3α increased during hospitalization, but only in patients with respiratory failure. No changes were observed for IFABP or CCL25 during hospitalization (Fig. 5B-C).

**Figure 5.**
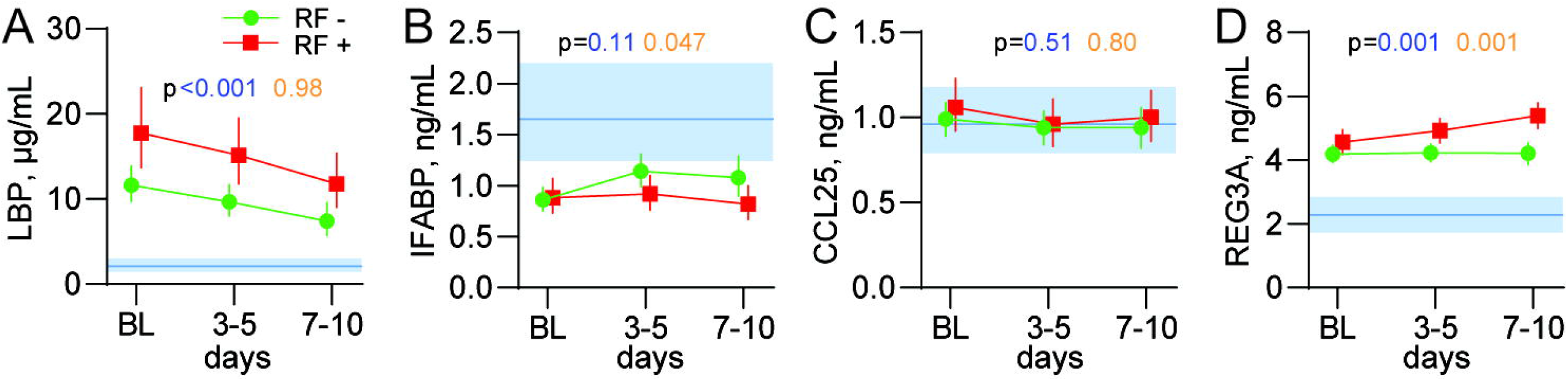
Temporal profile of markers of gut barrier dysfunction in relation to acute respiratory failure (RF) during hospitalization (P/F-ratio <26.6 kPa). A) LBP B) IFABP C) CCL25 and D) REG-3α. The yellow p-values reflect the effect RF from the repeated measures regression analysis, while the blue p-values reflect the interaction between time and RF. Blue areas reflect levels in age- and sex-matched healthy controls (n=24). Observations per time point: BL, n=144; 3-5 days, n=134; 7-10 days, n=84. Abbreviations: LBP, lipopolysaccharide-binding protein; IFABP, intestinal-type fatty acid-binding protein; CCL25, chemokine (C-C motif) ligand 25; REG-3α, regenerating islet-derived protein 3 alpha; RF, respiratory failure.

### The association of LBP with persistent respiratory dysfunction and inflammation

Notably, LBP levels after three months were higher in patients with respiratory dysfunction (DL_CO_<LLN) (Supplementary Figure 1), and negatively correlated with per cent predicted values of DL_CO_ (ρ -0.27, p=0.031). Of note, we found no correlation between LBP and measures of microbiota diversity in the subgroup with both available. Plasma levels of LBP at admission and after three months were significantly associated with several clinical markers of systemic inflammation, including CRP levels, white blood cell and neutrophil count (Supplementary Fig. 2), potentially relating gut leakage to persistent systemic inflammation even several months after hospitalization.

## Discussion

In this sub-study of the NOR-Solidarity trial, we investigated a potential gut-lung axis during and after hospitalization for COVID-19. Our results can be summarized as follows: i) three months after hospitalization for COVID-19, patients with persistent respiratory dysfunction (DL_CO_<LLN) showed a lower microbial diversity and an altered global gut microbiota composition than patients with normal respiratory function, ii) these microbiota alterations included reduced abundance of *Erysipelotrichaceae UCG-003* and increased abundances of *Veillonella* and *Flavonifractor*, iii) during hospitalization, increased plasma levels of LBP were strongly associated with acute respiratory failure, iv) LBP levels remained elevated during and after hospitalization, and were significantly associated with persistent low-grade inflammation and respiratory dysfunction at the three-month follow-up.

In a recent report, Yeoh *et al* reported downregulation of several gut commensals with known immunomodulatory potential in COVID-19 patients, including *Faecalibacterium prausnitzii* and *Eubacterium rectale*, which remained low in samples collected up to 30 days after disease resolution(10). Herein, we extend these findings by showing that altered gut microbiota, including decreased alpha diversity three months after hospital admission was associated with impaired pulmonary function at this time point. We also found that several members of the Lachnospiraceae and Ruminococcaceae families, such as *Coprococcus* and *Ruminococcus*, known producers of butyrate, were reduced in COVID-19 patients with pulmonary impairment after three months. Butyrate has local immunomodulatory effects in the gut mucosa, and as the main energy substrate for enterocytes, it is vital for gut barrier maintenance(29). Interestingly, LBP levels as an indirect marker of gut leakage were associated with impaired pulmonary function, not only during hospitalization (P/F-ratio <26.6 kPa; respiratory failure), but also after three months (inverse correlation with DL_CO_).

Of interest, the COVID-related dysbiosis reported by Yeoh *et al* correlated with several inflammatory markers, in line with our finding of LBP being associated with acute inflammation during hospitalization. Notably, we now show that LBP levels are associated with persistent low-grade inflammation even after three months. A potential role of gut barrier dysfunction as a driver for the multisystem inflammatory syndrome occurring in pediatric COVID-19 was recently reported(30), and our findings suggest that similar mechanisms could be relevant in adult patients. Whereas LBP levels showed a declining trend over time, the opposite was observed for REG-3α, which increased during hospitalization, but only in patients with respiratory failure. REG-3α is a bactericidal C-type lectin produced in the intestine, with antibacterial properties against Gram positive bacteria, and with a role in maintaining a functional barrier between the gut mucosa and the gut microbial content(31).

Hence, a potential interpretation of our findings could be that both elevated LBP levels and downregulation of REG-3α at admission might reflect an impaired gut barrier. In some chronic inflammatory conditions related to immunodeficiencies, we have also seen a direct correlation between microbiota composition and markers of gut barrier function(32, 33), but this was not seen in the COVID-19 patients at three months. To what degree gut microbiota alterations are directly influencing the gut barrier, is therefore uncertain.

Other microbiota related traits could also be relevant. The largest effect size observed in patients with persistent pulmonary dysfunction was the increased relative abundance of *Veillonella*, which has previously been linked to several disease states of the lung and liver where fibrosis is central to the pathogenesis(28, 34, 35). Interestingly, increased relative abundance of *Veillonella* in COVID-19 patients was also recently reported by Tao *et al*(36).However, whether this bacterial genus could be relevant for fibrosis development after COVID-19, either as a contributing factor or as a consequence of a fibrotic process, cannot be answered in an observational study.

Whether the observed alterations in microbiota composition and gut leakage markers in this trial are related to COVID-19 *per se* or mirror other factors, such as comorbidities and treatment, is not clear. In multivariate analyses, LBP was strongly associated with acute respiratory failure after adjustment for age, gender, treatment, comorbidities and use of antibiotics. LBP is an acute phase protein induced in the liver by several stimuli, and could reflect an acute phase reaction as well as a signal from the gut(37). Also, the fact that both LBP and global gut microbiota composition associated with persistent respiratory dysfunction, irrespective of antibiotics use during hospitalization, suggest an involvement of gut microbial environment.

The present study has some limitations. Due to logistic constraints of launching a randomized trial during the first wave of the pandemic, the gut microbiota samples were only collected at the three-month follow-up. We therefore cannot relate the long-term microbiota alterations to potential microbiota alterations during hospitalization. Also, the analysis of microbiota composition was only performed in one cohort, and without a validation panel these must be considered explorative. Moreover, pulmonary function tests beyond P/F-ratio were only performed at three-month follow-up, eliminating the possibility for baseline comparisons. The study also has obvious strengths, including standardized data capture in a randomized trial with longitudinal biobanking, as well as comprehensive long-term follow-up with blood tests, microbiota sampling and assessment of pulmonary function by DL_CO_. Also, to our knowledge, our study is the first to link pulmonary function to gut microbiota alterations in COVID-19.

In conclusion, the decreased microbial diversity and compositional gut microbiota alterations in patients with persistent respiratory dysfunction, as well as the association of persistently raised LBP levels with these clinical features, point to a potential gut-lung axis in COVID-19. These observations could be related to not only acute respiratory failure during hospitalization, but also to long-term COVID-19 morbidity. Our findings warrant further research on the potential role of gut microbiota composition and gut barrier dysfunction in relation to long-term pulmonary dysfunction and long COVID.

## Data Availability

Data from the present study are available from the corresponding author on reasonable request.

## Acknowledgements

We thank WHO Solidarity and NOR-Solidarity study groups for the opportunity to perform this add-on study. We would also like to thank Karoline Hansen Skåra and Azita Rashidi at the Institute of Internal Medicine, Oslo University Hospital, Rikshospitalet, for their contributions to the biobank collection, and Mona Skjelland for access to the control plasma biobank. Moreover, we thank Fredrik Müller and Cahtrine Fladeby at the Department of Microbiology, Oslo University Hospital, for access to the viral analysis data.

## Conflicts of interest

All authors declare no competing interest.

## Funding

The study was funded by the National Clinical Therapy Research in the Specialist Health Services (KLINBEFORSK), Norway and South-Eastern Norway Regional Health Authority (grant number 2021071). The microbiota analyses were funded by the strategic research area “Personalized microbiota therapy in clinical medicine” at Oslo University Hospital. The funders had no role in study design, data collection, data analysis, data interpretation, or writing of the report. The corresponding author had full access to all the data in the study and had final responsibility for the decision to submit for publication.

## Contributions

BV, TU, PA, JRH and MT were responsible for the study conception and execution of the present substudy. ABD, TK, ICO, AMDR, KNH, PA and MT were responsible for the management, coordination, research activity planning and execution of the NOR-Solidarity trial. TL and OHS were responsible for the three-month follow-up protocol for pulmonary function. TBD, ABD, BH, TR and PA coordinated the collection and storage of the biobank material. AMDR, BS, KT, HH, AT, RM, MH, RE, ÅB, ABK, AJ, LT, HS, BRK, RBO, CMY, NVS, RH, SA and BB were locally responsible for conducting the trial at the various included hospitals providing rectal swab material. BV performed the DNA extraction and library preparation of microbiota samples. TU, TL and AEM performed the gut leakage marker analyses. BV and TU performed the statistical analyses. KH performed the bioinformatics analyses of microbiota data. BV and MT drafted the manuscript. TU, ABD, PA and JRH critically revised the manuscript. All authors revised and approved the final version of the manuscript.

## Notes

### Competing Interest Statement

The authors have declared no competing interest.

### Clinical Trial

NCT04321616

### Author Declarations

The study was approved by the Committee for Medical Research Ethics Region South East Norway (118684) and by the Norwegian Medicines Agency (20/04950-23).

