## Supplemental material for "Gut microbiota alterations in patients with persistent respiratory dysfunction three months after severe COVID-19"

### Supplementary material

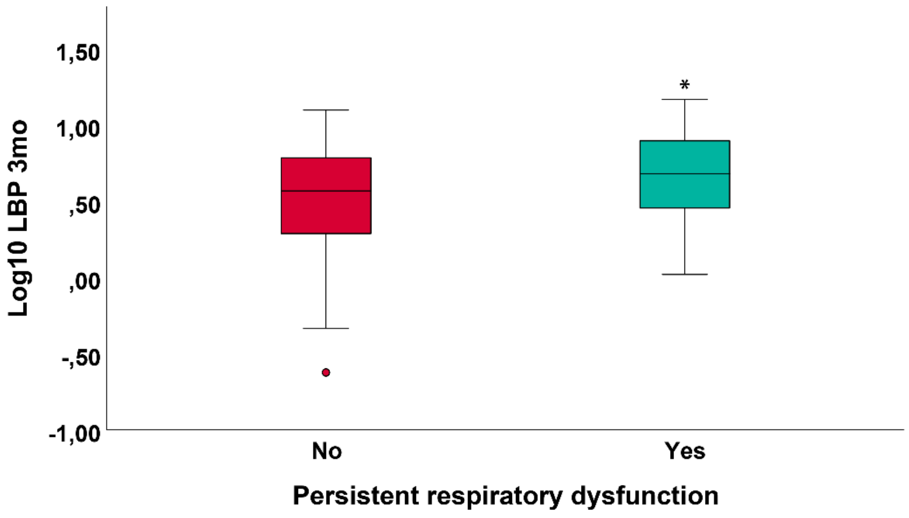


**Supplementary Figure 1.** Levels of LBP measured at the three-month follow-up in patients with or without persistent respiratory dysfunction ($DL_{CO}$<LLN). *p<0.05, 2-tailed Student's t-test. LBP, lipopolysaccharide-binding protein.

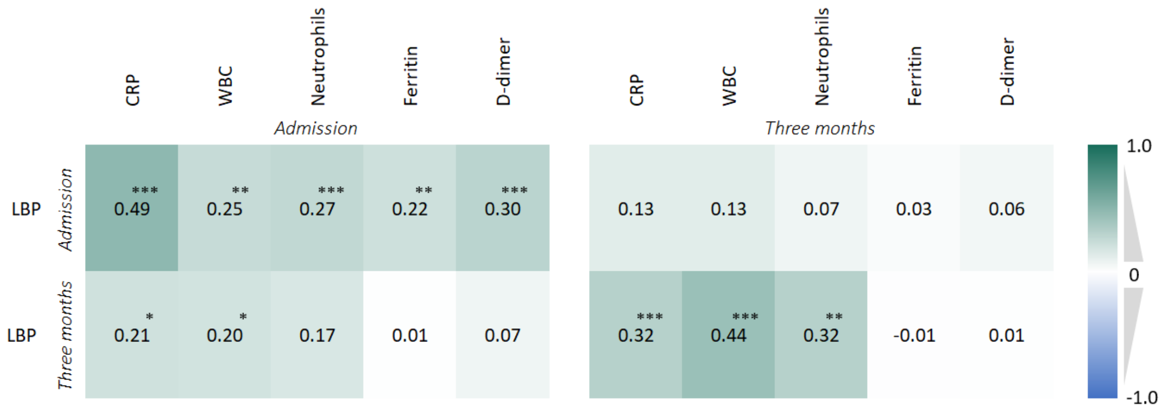


**Supplementary Figure 2.** Correlation heatmap of LBP levels with inflammation markers at corresponding time points. Values are Spearman's ρ with *p<0.05, **p<0.01, ***p≤0.001. Abbreviations: LBP, lipopolysaccharide-binding protein; CRP, C-reactive protein; WBC, white blood cell count.
